# Who Develops Pandemic Fatigue?

**DOI:** 10.1101/2022.01.24.22269786

**Authors:** Steven Taylor, Geoffrey S. Rachor, Gordon J. G. Asmundson

**Affiliations:** Department of Psychiatry, University of British Columbia, Vancouver, BC, CANADA; Department of Psychology, University of Regina, Regina, SK, CANADA

## Abstract

According to the World Health Organization, pandemic fatigue poses a serious threat for managing COVID-19. The cardinal feature of pandemic fatigue is a progressive decline in adherence to social distancing (SDIS) guidelines, which is associated with pandemic-related emotional burnout. Little is known about the nature of pandemic fatigue; for example, it is unclear who is most likely to develop pandemic fatigue. We sought to evaluate this issue based on data from 5,812 American and Canadian adults recruited during the second year of the COVID-19 pandemic. Past-year decline in adherence to SDIS had a categorical latent structure according to Latent Class Analysis, consisting of an SDIS adherent group (Class 1: 92% of the sample) and a group reporting a progressive decline in adherence to SDIS (i.e., pandemic fatigue; Class 2: 8% of the sample). Class 2, compared to Class 1, was associated with greater pandemic-related burnout, pessimism, and apathy about the COVID-19 pandemic. They also tended to be younger, perceived themselves to be more affluent, tended to have greater levels of narcissism, entitlement, and gregariousness, and were more likely to report having been previously infected with SARSCOV2, which they regarded as an exaggerated threat. People in Class 2 also self-reported higher levels of pandemic-related stress, anxiety, and depression, and described making active efforts at coping with SDIS restrictions that they perceived as unnecessary and stressful. People in Class 1 generally reported that they engaged in SDIS for the benefit of themselves and their community, although 35% of this class also feared they would be publicly shamed if they did not comply with SDIS guidelines. The findings suggest that pandemic fatigue affects a substantial minority of people and even many SDIS-adherent people experience emotionally adverse effects (i.e., fear of being shamed). Implications for the future of SDIS are discussed.

## Introduction

According to the World Health Organization (WHO), pandemic fatigue during the current COVID-19 crisis is a global problem that “poses a serious threat to efforts to control the spread of the virus” (p. 6) (1). The cardinal feature of pandemic fatigue is a progressive decline in adherence to government guidelines for social distancing (SDIS), arising in the weeks or months in which SDIS and other pandemic-mitigation restrictions are in place. The decline in adherence is hypothesized to be associated with pandemic-related burnout (1), which involves cynical or negative attitudes about the nature and origins of COVID-19 (e.g., belief that the threat of COVID-19 is exaggerated or a hoax), and pessimism, apathy, or hopelessness about methods for reducing the pandemic, such as SDIS. Pandemic fatigue is a chronic stress reaction in which the response to the stressor (i.e., decline in adherence to SDIS) perpetuates the stressor (i.e., facilitates the spread of pandemic infection) (2). As pandemic fatigue sets in, people become increasingly lax about staying safe from infection; specifically, they increasingly disregard SDIS guidelines and may defy public health guidelines by holding covert social gatherings or taking clandestine trips abroad (1-4).

A 2020 survey of over 7,000 American adults during the COVID-19 pandemic found that pandemic fatigue was characterized by progressively worsening adherence to the following SDIS guidelines: (a) remaining in residence except for essential activities or exercise, (b) having no close contact with non-household members, (c) not having visitors over to one’s home, and (d) avoiding eating at restaurants (5). Adherence to mask wearing showed little or no decline over time (5), possibly because masks were mandated in many public places (e.g., stores, public transit), whereas it was more difficult for authorities to regulate social activities such as having visitors to one’s home. Other studies have reported similar findings in numerous countries (6), although it is unclear whether some individuals are more prone to pandemic fatigue than other people. Pandemic fatigue is not unique to COVID-19. Similar phenomena have been described in past pandemics. For example, during the 1918 Spanish flu pandemic, public cooperation with SDIS mandates deteriorated with successive waves of infection (7). During both COVID-19 and the Spanish flu, people in many communities objected to widespread closures and wanted to lift restrictions and resume normal life, despite active, widespread infection (8-10).

Declining motivation to adhere to SDIS may be due to range of psychological and other factors, including decreases in perceived risk as people become habituated to their changed lifestyles during the COVID-19 pandemic. Other factors potentially contributing to pandemic fatigue include accumulating costs or hardships, such as growing economic losses, difficulties working or studying from home, and social isolation arising from business closures, stay-at-home orders, and other social and occupational restrictions associated with SDIS mandates (1). Previous studies during the COVID-19 pandemic found that non-adherence to lockdown was associated with the perception that lockdown is unnecessary and ineffective (11), with younger age (12-15), greater perceived personal affluence (16), and lower trust in government (16). Gender findings have been mixed, with some studies finding that non-adherence is more prevalent in men (12, 16) and others reporting greater prevalence in women (17).

Much remains to be learned about the nature of pandemic fatigue. It is unclear whether the decline in SDIS is unifactorial; that is, do all forms of SDIS progressively decline over time or are some forms of SDIS more likely to be adhered to than others? If non-adherence to SDIS is unifactorial, then the question arises as to whether this factor has a dimensional or categorical structure; that is, pandemic fatigue might be a matter of degree or, alternatively, there might be distinct types of people, such as those who generally adhere to SDIS versus those who progressively become increasingly non-adherent. Also, little is known about correlates of declining adherence to SDIS. Although declining adherence may be correlated with burnout, it may also be correlated with a range of variables such as anxiety, depression, and loneliness due to SDIS. The relationship between declining adherence and personality traits also remains to be investigated. Identifying the correlates of declining adherence can provide insights as who is most likely to develop pandemic fatigue. Identifying at-risk groups for non-adherence can help to guide efforts at reinvigorating communities to follow SDIS protocols (1).

The aims of the present study were to (a) investigate the structure and correlates of declining adherence to pandemic-related SDIS guidelines, based on data collected during the COVID-19 pandemic, (b) to identify the demographic, cognitive, affective, and personality characteristics of people most likely to become progressively non-adherent, and (c) to investigate the reasons why other people adhered to SDIS guidelines. At the time of conducting the study, social distancing was in place along with closures or restricted operations of restaurants, and restrictions against travel and against large social gatherings.

In the present study, cognitive and affective characteristics of COVID-19-related burnout were assessed broadly, including COVID-19-related burnout and associated features such as apathy, pessimism, blame, distrust in government efforts to stem the pandemic, along with beliefs that the COVID-19 threat is exaggerated and conspiratorial beliefs that COVID-19 is a hoax. People who disregard SDIS are also more likely to have been infected with the coronavirus causing COVID-19, and they may experience only mild symptoms, as commonly occurs in COVID-19 (18), which can reinforce beliefs that the COVID-19 threat is exaggerated, thereby amplifying non-adherence. Accordingly, we assessed whether respondents believed that they had acquired COVID-19. Belief that one had been infected, rather than objective evidence of infection, is important because beliefs drive behaviors. Anxiety, depression, stressors, and coping strategies were also assessed, as stressors and distress are thought to exacerbate non-adherence to SDIS (1). People who are especially distressed may be most likely to violate SDIS guidelines in pursuit of socially rewarding activities, such as attending social gatherings. To investigate the relationship between pandemic fatigue and personality, broad and narrow personality traits were assessed. Broad traits consisted of the Big 5 (i.e., agreeableness, conscientiousness, negative emotionality, extraversion, and openness to experience) (19). Adherence to SDIS requires that people put the well-being of the community ahead of personal self-interests. Accordingly, people who score highly on personality traits such as narcissism or self-entitlement might be most likely to be non-adherent to SDIS, particularly when SDIS guidelines are personally inconvenient (e.g., refraining from socializing).

Narcissism and psychological entitlement are related but distinguishable constructs. Entitlement refers to a stable and pervasive sense that one deserves more compared to other people (34). Narcissism is a broader construct involving self-absorption, grandiosity, arrogance, and a sense of entitlement (53). Thus, entitlement can be a component of narcissism but high levels of entitlement can also occur in the absence of narcissism; that is, a person can feel entitled to special treatment without necessarily having an inflated sense of self-worth. A sense of relative deprivation is one way in a person might feel entitled without necessarily being grandiose; for example, “I was deprived of my grad party last year because of lockdown, so I deserve to be having fun with my friends this year.”

## Materials and Methods

### Sample

The sample consisted of 5,812 adults (age ≥18 years) from the U.S. (*n*=2,964) and Canada (*n*=2,848) who were recruited as part of the COVID Stress Study (20, 21), which is a broad investigation into the psychology of COVID-19. The mean age of the sample was 49 years (SD=17 years, range 18-92 years). About half the sample (52%) were employed full- or part-time, most (78%) had completed full or partial college, and 57% were female. Most (64%) were White, with the remainder being Asian (13%), African American/Black (11%), Latino/Hispanic (4%), or other (7%). A total of 4% of the sample reported that they were healthcare workers and 7% stated that they had been diagnosed with COVID-19 by a healthcare worker. A third (32%) of respondents reported that they had been partially or fully vaccinated against the novel coronavirus at the time of the study, and 38% reported that they had a preexisting (pre-COVID-19) general medical condition. A total of 23% of respondents reported that they had a recent (past year) history of a mental health problems, predominantly mood or anxiety symptoms, and 71% believed that COVID-19 had harmed their mental health.

### Measures

The respondent’s perceived socioeconomic status, in relation to people in one’s country, was assessed by a 10-point item from the MacArthur Scale of Subjective Social Status, which has been shown to have good reliability and validity (22). Higher scores corresponded to greater perceived socioeconomic status. Political conservatism was assessed with a face-valid item, “In general, how would you describe your political views?” (1 = very liberal, 7 = very conservative). The remaining measures, listed in Table 1, were multi-item scales. Table 1 shows the internal consistency reliabilities for the multi-item scales. McDonald’s ω total (23), which is a commonly used alternative to Cronbach’s α, was used as the measure of reliability. McDonald’s ω was used instead of Cronbach’s α because the latter tends to underestimate reliability (24). Values of ω are interpreted in the same way as α; that is, values in the range of 0.70-0.80 indicate acceptable reliability, 0.80-0.90 are good, and values greater than 0.90 are excellent. Table 1 shows that all scales had at least acceptable reliability, and almost all (93%) had good-to-excellent reliability.

**Table 1.** Internal consistency reliability coefficients for multi-item scales.

| Scale | No. items | $\omega$ total |
| --- | --- | --- |
| Agreeableness | 2 | 0.76 |
| Anxiety | 7 | 0.96 |
| Belief in COVID-19 conspiracy theories | 4 | 0.92 |
| Conscientiousness | 2 | 0.82 |
| Coping with COVID-19 | 38 | 0.94 |
| COVID-19 anti-vaccination attitudes, current | 12 | 0.93 |
| COVID-19 apathy | 7 | 0.89 |
| COVID-19 blame | 8 | 0.88 |
| COVID-19 burnout | 12 | 0.96 |
| COVID-19 exaggerated | 3 | 0.90 |
| COVID-19 pessimism | 16 | 0.93 |
| COVID-19 threat is exaggerated | 3 | 0.90 |
| Depression | 9 | 0.94 |
| Disregard for social distancing, current | 4 | 0.90 |
| Distrust in government | 14 | 0.93 |
| Entitlement | 12 | 0.90 |
| Extraversion | 2 | 0.84 |
| Facemask non-adherence, current | 3 | 0.86 |
| Loneliness | 3 | 0.91 |
| Narcissism | 7 | 0.89 |
| Negative emotionality | 2 | 0.87 |
| Openness to experience | 2 | 0.76 |
| Reasons for social distancing | 8 | 0.86 |
| Robust personal health | 3 | 0.91 |
| SDIS: Changes in adherence to social distancing | 10 | 0.87 |
| Sociability | 5 | 0.90 |
| Stressors associated with COVID-19 | 22 | 0.95 |

Past-year changes in adherence to SDIS were assessed by the 10-item face-valid SDIS Scale, developed for the purpose of the present study. The items, listed in Table 2, assessed respondent’s reports of whether their adherence to SDIS had increased, remained unchanged, or decreased over the past year. For people who adhered to SDIS, their reasons for adherence were assessed by an 8-item face-valid scale in which respondents rated their strength of agreement with each item. The items appear in Table 3.

**Table 2.** Endorsement and factor loadings for items assessing changes in adherence to social distancing over the past year (total sample).

| Item | % less often | % no change | % more often* | Factor loading |
| --- | --- | --- | --- | --- |
| Close contact (within 6 feet) with people who don't live with me | 44 | 45 | 12 | 0.50 |
| Go out to socialize with friends, family, or relatives | 54 | 37 | 9 | 0.66 |
| Have friends, neighbors, or relatives to my home | 48 | 44 | 8 | 0.64 |
| Socialize outside of my social bubble | 48 | 46 | 6 | 0.62 |
| Go to gatherings of more than 6 people | 46 | 49 | 6 | 0.65 |
| Have contact with people who could be high-risk for COVID-19 | 39 | 57 | 4 | 0.50 |
| Go to crowded public places where social distancing is not possible | 45 | 51 | 4 | 0.60 |
| Go to a crowded bar, club, restaurant, or cafe | 42 | 55 | 4 | 0.57 |
| Travel outside of my state or province | 42 | 55 | 4 | 0.50 |
| Go to crowded church services | 31 | 66 | 3 | 0.50 |

\*Indicates progressive decline in adherence to social distancing (SDIS).

**Table 3.**
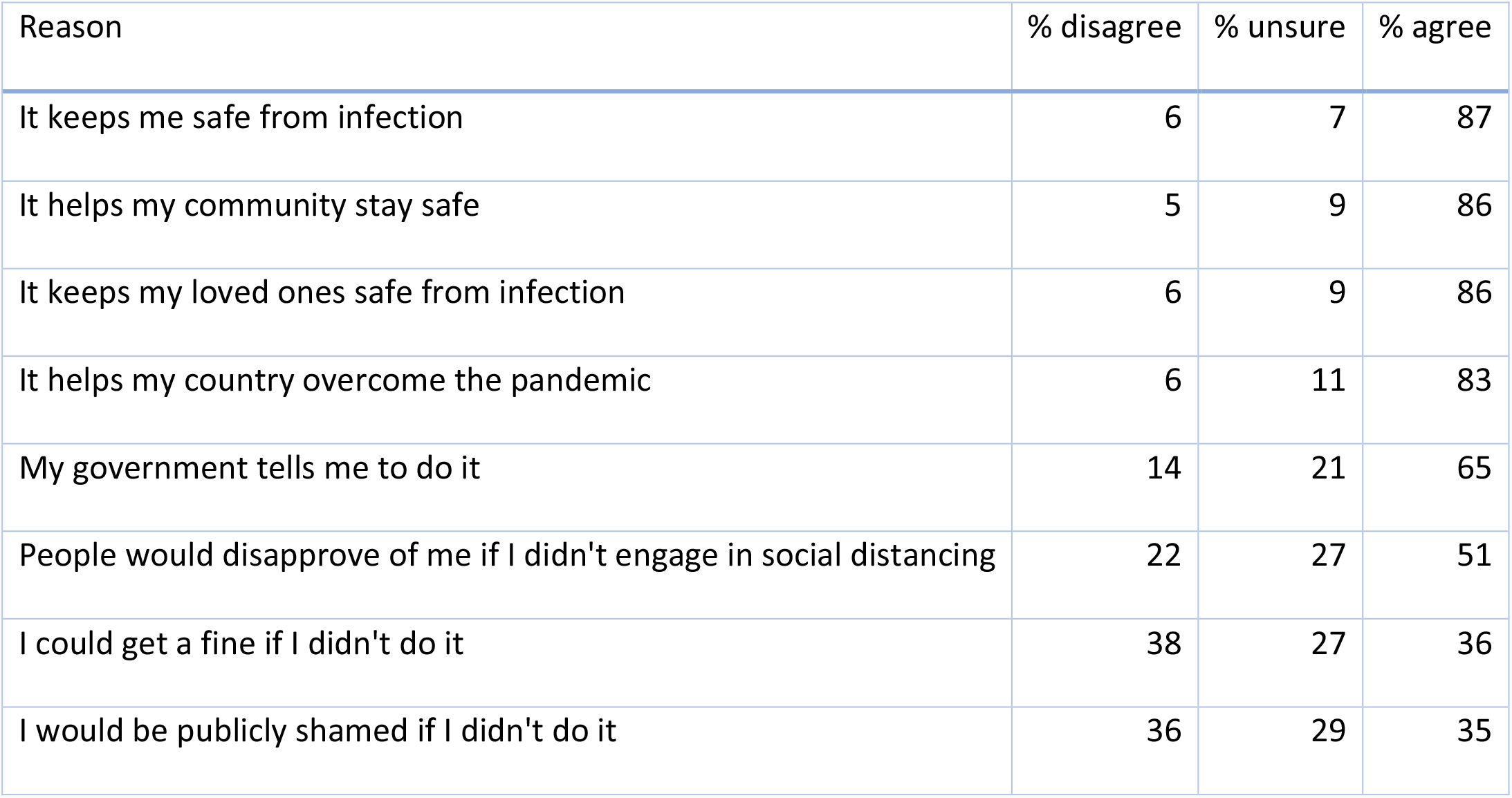
Reasons for adhering to social distancing guidelines (Class 2 participants).

Scales developed in our previous research (25) were administered to assess the following: (a) current disregard for SDIS (as distinct from past-year changes in SDIS), (b) belief that the dangerousness of COVID-19 is exaggerated, (c) belief that one has robust personal health against infection, and (d) belief in COVID-19 related conspiracy theories. Items on these measures were rated on a 5-point scale (0=strongly disagree, 4=strongly agree). These face-valid scales have good levels of reliability and validity (25) (see Table 1).

COVID-19 related stressors were assessed using a 22-item face-valid scale based on our previous research (21) in which respondents rated the frequency (1=never, 5=often) of various stressors experienced during COVID-19 (e.g., difficulty working from home, isolation, crowding at home, difficulty caring for loved ones). Despite covering a range of different, commonly occurring stressors during COVID-19 (21), the 22-item scale had excellent reliability (see Table 1). Coping with COVID-19-related stress was assessed with a 38-item, face valid scale (21) that assessed the frequency of use (1=never, 5=very often) of a range of different coping strategies (e.g., exercising, reading novels, talking with a trusted friend). The reliability of the scale was excellent (see Table 1).

Current anti-vaccination attitudes toward COVID-19 vaccines were measured using an adaptation of the Vaccination Attitudes Examination Scale (26), assessing vaccination attitudes specific to COVID-19 (27). The items in this scale, each rated on a 6-point scale (0=strongly disagree, 5=strongly agree), assess mistrust of vaccine benefit, worries over unforeseen future effects of the vaccine, concerns about commercial profiteering from the vaccine, and preference for natural immunity. The scale has good levels of reliability and validity (26, 27) (see Table 1). Current non-adherence for wearing facemasks was assessed by a 3-item face-valid scale in which respondents were asked to rate the frequency in which they intentionally refrained from wearing a mask in public places (e.g., in stores or on public transit). Items were rated on a 1-6 scale (0=never, 6=more than once a day).

COVID-19-related burnout, apathy, blame, and pessimism were assessed using scales developed for the present study. All had good reliability (see Table 1). These measures were based on the short form of the Burnout Measure (28), which is a psychometrically sound measure of burnout. In the COVID-19-related burnout scale, respondents were presented with 12 adjective statements (e.g., irritable, frustrated, emotionally exhausted) and were asked, “When you think about COVID-19, how often do you feel the following?” Statements were rated on a 5-point scale (1=never, 5=always). COVID-19-related apathy was measured by 7 statements, each rated on a 5-point scale (e.g., “Regardless of what we do, almost everyone will get COVID-19”; 1=strongly disagree, 5=strongly agree). The same rating scale was used in the COVID-19-related blame and pessimism scales. Blame was assessed by 8 items (e.g., “People in my community are to blame for the spread of COVID-19”). COVID-19-related pessimism was assessed in a similar manner with 16 items (e.g., “There is nothing I can do to keep myself safe from COVID-19”). General anxiety and depression over the past week were assessed, respectively, by the GAD-7 (29) and PHQ-9 (30). Both scales have good psychometric properties (29, 30) (see Table 1).

A broad assessment of personality traits was conducted using the Ten Item Personality Inventory (TIPI) (19). The TIPI is a 10-item measure of the Big 5 personality dimensions of extraversion, agreeableness, conscientiousness, negative emotionality, and openness to experience. Despite being a very brief measure, the TIPI has performed well on various indices of reliability and validity (19, 31, 32). In the present study, the TIPI scales had acceptable-to-good levels of reliability (see Table 1).

Narcissism was measured using the 7-item scale from the Short Dark Tetrad (33), in which respondents rated their strength of agreement on a 5-point scale (e.g., “I have some exceptional qualities”; 1=strongly disagree, 5=strongly agree). This scale has good reliability and validity (33) (see Table 1). Psychological entitlement was assessed using a 12-item version of the Psychological Entitlement Scale (34), in which participants rated, on a 7-point scale, the extent to which the respondent believed that he or she was entitled to special treatment in various aspects of life (e.g., “I honestly feel I’m just more deserving than others”; 1=strongly disagree, 7=strongly agree). Although entitlement is related to narcissism (*r*=0.44 in the present study), the two constructs are distinguishable in that entitlement entails beliefs in deserving special treatment without necessarily entailing, as in narcissism, an inflated sense of self-worth. The scale has good reliability and validity (34) (see Table 1).

Sociability was assessed using the Sociability Scale (35), in which respondents rated their agreement on five statements (e.g., “I find people more stimulating than anything else”; 1=strongly disagree, 5=strongly agree). The scale has sound psychometric properties (35) (see Table 1). The tendency to feel lonely was measured using the Loneliness Scale (36) in which items assessing loneliness (e.g., “How often do you feel left out?”) are rated on a 3-point scale (1=hardly ever, 3=often). The scale has good psychometric properties (36) (see Table 1).

Distrust in government for managing the COVID-19 pandemic was assessed using a 14-item face-valid measure developed for the purpose of the present study. For each item, participants rated their agreement on a 5-point scale for statements such as “My government has allowed its citizens to be financially ruined by the pandemic” (1=strongly disagree, 5=strongly agree). The reliability of the scale was excellent (see Table 1).

### Data Collection Procedures

Data were collected from March 24 to May 4, 2021, at which time social distancing restrictions were implemented throughout the U.S. and Canada. The sample was obtained using an internet-based self-report survey delivered in English by Qualtrics, a commercial survey sampling and administration company. Qualtrics solicited the present sample, for which no data have yet been reported on or published, as part of our ongoing research program (20, 21, 37). Qualtrics maintains a pool of potential participants who have agreed to be contacted in order to respond to surveys. Qualtrics selected and contacted participants to meet sampling quotas to approximate general population demographics, based on age, gender, ethnicity, socioeconomic status, and geographic region within each country. The demographic composition of the sample approximated census-derived data of U.S. and Canadian adults (i.e., excluding children and adolescents), where, for example, the mean age averaged across countries is 50 years and 67% white. The sample departed from census data in that females were over-represented (57%) as compared to census data (51%). However, gender was not substantively associated with any of the variables in this study; that is, effect sizes for gender were smaller than what is conventionally regarded as “small” effect sizes (see below).

All respondents provided informed consent prior to completing the survey. The research described in this article was approved by the Research Ethics Board of the University of Regina (REB# 2020-043). Filters were used to eliminate data from careless responders. Embedded in the assessment battery were four attention-check items (e.g., “This is an attention check, please select Strongly Agree”; “For our research, it is really important that you paid attention while responding to our survey. How attentive were you when responding?”: “Very Inattentive” to “Very Attentive”). Participants were included only if they provided correct responses to three or more of the four attention checks (e.g., “Strongly agree” or “Very attentive”), indicating that they were sufficiently attentive. In addition, at the end of the assessment battery, participants were asked to indicate whether, in their honest opinion, their data should be used. Those who responded “no” were excluded from data analysis, regardless of their score on the attention-check items.

### Statistical Procedures

Exploratory factor analysis using robust Maximum-Likelihood and Parallel Analysis were used to determine the number of factors of the SDIS scale. This was followed by Latent Class Analyses, also using robust Maximum-Likelihood, to determine whether the factor was dimensional or categorical in nature. Analyses were conducted using SPSS 27.0 and Mplus (38). In the Latent Class Analyses, models consisting of increasing numbers of classes were evaluated (e.g., 1 vs 2 classes, 2 vs 3 classes) until the best-fitting model was identified, as determined by four goodness-of-fit indices: Akaike Information Criterion, Bayesian Information Criterion, sample-size adjusted Bayesian Information Criterion, and the Bootstrap Likelihood Ratio Test. For the first three fit indices, the best-fitting model has the lowest value on these indices. For the Bootstrap Likelihood Ratio Test, the best fitting model is a model consisting of N classes, which has a significantly better (*p*<0.01) fit than a model consisting of *N*−1 classes, and is not significantly different from a model consisting of *N*+1 classes. The resulting number of classes were then compared on a range of affective and other variables.

Given the number of analyses reported in this article, the α level was set at 0.01 instead of 0.05. This adjustment corrects for inflated Type I error without unduly inflating Type II error with a more stringent correction, such as a Bonferroni correction. Given the large sample size, substantively trivial effect sizes would be statistically significant (e.g., for *r*=0.05, *p*<.001). Accordingly, to facilitate the interpretation of correlations, we used Cohen’s criteria (39) to classify effect sizes as small, medium, or large. Effect sizes were either Cohen’s *d* for pairs of variables in which one or both variables were continuous, or Cramér’s *v* for comparisons involving pairs of nominal variables. Effect sizes for Cohen’s *d* are conventionally classified as small (*d*=0.20), medium (*d*=0.50), and large (*d*=0.80) (39). To give precision to these classifications for values of *d* falling between these values, we classified *d* in terms of ranges, using the midpoint between 0.20 and 0.50, and midpoint between 0.50 and 0.80, so as to distinguish among small, medium, and large values of *d*; that is, small 0.20–0.349, medium 0.35–0.649, and large >0.65. The corresponding criteria for classifying Cramér’s *v* and the range for interpreting scores were as follows: small (0.1, 0.1-0.19), medium (0.3, 0.2-0.39), and large (0.5, ≥ 0.40).

## Results

Exploratory factor analysis of the SDIS items indicated a single-factor solution with only the first Eigen value being greater than 1.00. The first 5 Eigen values were 3.98, 0.98, 0.80, 0.74, and 0.69. Factor loadings are shown in Table 2, indicating that all loadings were salient (>0.30). The SDIS scale, representing the sum of the 10 SDIS items, had a high internal consistency (Table 1). The total score was used as the input variable for the Latent Class Analyses. The results, shown in Table 4, indicated that the best fitting model consisted of two classes; Class 1 (92% of sample, *n*=5,326), Class 2 (8%, *n*=486). The results show that the majority of participants were adherent to SDIS; but, a sizeable minority (8%) reported a deterioration in SDIS, indicative of pandemic fatigue.

**Table 4.** Fit indices for latent class analysis.

| No. classes | Akaike Information Criterion | Bayesian Information Criterion | Sample size adjusted Bayesian Information Criterion | Bootstrap likelihood ratio difference test $\chi^2(df = 2)$ | Bootstrap likelihood ratio difference test $p$ |
| --- | --- | --- | --- | --- | --- |
| 1 | 20,761.93 | 20,775.26 | 20,768.91 | -- | -- |
| 2 | **15,878.47** | **15,905.15** | **15,892.43** | **4,887.45** | < 0.001 |
| 3 | 15,882.47 | 15,922.48 | 15,903.41 | < 0.01 | > 0.999 |

*Note.* Bold = best-fitting model; -- = not applicable.

Table 5 shows the comparisons between classes on a range of personality, affective, and COVID-19-related variables. Statistical power to detect small effect sizes at α=0.01 was 0.95 for these analyses. Accordingly, the study was sufficiently powered to detect even small differences between classes. To facilitate the interpretation of the results, a Discriminant Function Analysis was conducted to determine which variables best distinguished the two classes. Input variables for this analysis were those having small, medium, or large effects in Table 5. The table shows the loadings of the variables on the discriminant function. Salient (>0.30) loadings are in bold. Taking both the effect sizes and discriminant loadings into consideration, Table 5 shows that people in Class 2, compared to Class 1, reported a greater current disregard for SDIS. In other words, for Class 2 there was a progressive past-year deterioration in SDIS as well as a current low level of adherence.

**Table 5.** Comparisons between classes.

| Variable | Class 1:<br>M (SD)<br>or % | Class 2:<br>M (SD)<br>or % | $t(df=5,810)$<br>or<br>$\chi(df=1)$ | $p$ | ES | Type<br>of ES | DFA<br>loading |
| --- | --- | --- | --- | --- | --- | --- | --- |
| Disregard for SDIS, current | 2.2 (3.1) | 5.2 (4.7) | 19.76 | < 0.001 | 0.94 | *d* | **0.60** |
| Coping with COVID-19 | 90.4 (24.1) | 113.1 (30.5) | 19.44 | < 0.001 | 0.92 | *d* | **0.59** |
| Stressors associated with COVID-19 | 40.4 (15.3) | 54.0 (21.8) | 18.07 | < 0.001 | 0.86 | *d* | **0.54** |
| Belief in COVID-19 conspiracy theories | 2.2 (3.2) | 4.9 (4.7) | 17.42 | < 0.001 | 0.83 | *d* | **0.53** |
| COVID-19 exaggerated | 3.1 (3.1) | 5.4 (3.7) | 15.14 | < 0.001 | 0.72 | *d* | **0.46** |
| COVID-19 apathy | 16.2 (5.6) | 19.8 (7.0) | 13.12 | < 0.001 | 0.62 | *d* | **0.40** |
| Robust personal health | 4.5 (2.9) | 6.2 (3.1) | 12.34 | < 0.001 | 0.59 | *d* | **0.37** |
| Narcissism | 20.2 (5.8) | 23.5 (5.8) | 12.14 | < 0.001 | 0.58 | *d* | **0.37** |
| Age (years) | 49.3 (17.0) | 40.8 (15.2) | 10.70 | < 0.001 | 0.51 | *d* | **-0.32** |
| Perceived socioeconomic status | 5.7 (1.9) | 6.6 (1.9) | 10.19 | < 0.001 | 0.48 | *d* | **0.31** |
| Anxiety | 4.7 (5.6) | 7.3 (6.0) | 9.66 | < 0.001 | 0.46 | *d* | 0.29 |
| Depression | 5.9 (6.7) | 9.0 (7.6) | 9.57 | < 0.001 | 0.45 | *d* | 0.29 |
| Sociability | 16.8 (4.6) | 18.7 (3.9) | 8.84 | < 0.001 | 0.42 | *d* | 0.27 |
| COVID-19 anti-vaccination attitudes, current | 36.6 (12.9) | 41.6 (12.9) | 8.15 | < 0.001 | 0.39 | *d* | 0.25 |
| COVID-19 pessimism | 38.1 (11.6) | 42.6 (14.8) | 7.96 | < 0.001 | 0.38 | *d* | 0.24 |
| Entitlement | 3.6 (1.1) | 4.0 (1.3) | 7.72 | < 0.001 | 0.37 | *d* | 0.23 |
| Agreeableness | 10.4 (2.3) | 9.7 (2.3) | 7.12 | < 0.001 | 0.34 | *d* | -0.21 |
| Conscientiousness | 11.0 (2.4) | 10.3 (2.6) | 5.76 | < 0.001 | 0.27 | *d* | -0.17 |
| Extraversion | 7.3 (3.0) | 8.0 (2.6) | 5.17 | < 0.001 | 0.25 | *d* | 0.16 |
| COVID-19 burnout | 29.1 (12.4) | 31.8 (12.4) | 4.74 | < 0.001 | 0.23 | *d* | 0.14 |
| Self-reported diagnosis of COVID-19 | 7 | 28 | 264.37 | < 0.001 | 0.21 | *v* | **0.50** |
| Belief that COVID-19 has harmed one's mental health | 0.7 (0.5) | 0.8 (0.4) | 3.95 | < 0.001 | 0.19 | *d* | -- |
| Country (1=U.S., 0=Canada) | 49 | 77 | 137.75 | < 0.001 | 0.15 | *v* | **0.36** |
| Loneliness | 5.3 (1.9) | 5.6 (1.9) | 2.78 | 0.005 | 0.13 | *d* | -- |
| Political conservatism | 3.6 (1.7) | 3.8 (1.9) | 1.64 | 0.101 | 0.08 | *d* | -- |
| Negative emotionality | 7.0 (1.0) | 7.1 (1.0) | 1.63 | 0.102 | 0.08 | *d* | -- |
| Openness to Experience | 9.9 (2.4) | 9.7 (2.4) | 1.67 | 0.095 | 0.08 | *d* | -- |
| Current mental health diagnosis | 23 | 26 | 3.28 | 0.070 | 0.07 | *v* | -- |
| Distrust in government | 0.7 (12.2) | 1.4 (10.8) | 1.19 | 0.234 | 0.06 | *d* | -- |
| Personally knew someone diagnosed with COVID-19 | 53 | 64 | 21.52 | < 0.001 | 0.06 | *v* | -- |
| Prior medical condition | 38 | 28 | 22.19 | < 0.001 | 0.06 | *v* | -- |
| Female (1=female, 0=other) | 58 | 48 | 17.17 | < 0.001 | 0.05 | *v* | -- |
| College (1=full or partial college, 0=less than college) | 78 | 85 | 16.40 | < 0.001 | 0.05 | *v* | -- |
| Received one or both doses of COVID-19 vaccine | 32 | 41 | 15.46 | < 0.001 | 0.05 | *v* | -- |
| Healthcare worker | 4 | 8 | 13.16 | < 0.001 | 0.05 | *v* | -- |
| Unemployed (1=unemployed, 0=other) | 9 | 5 | 7.97 | 0.005 | 0.04 | *v* | -- |
| Facemask non-adherence, current | 13.1 (3.6) | 13.0 (3.2) | 0.77 | 0.440 | 0.04 | *d* | -- |
| Single (1=single, 0=other marital status) | 47 | 41 | 5.73 | 0.017 | 0.03 | *v* | -- |
| White (1=White, 0=other) | 65 | 59 | 6.84 | 0.009 | 0.03 | *v* | -- |
| COVID-19 blame | 23.4 (6.8) | 23.5 (7.5) | 0.48 | 0.634 | 0.02 | *d* | -- |

$d$ = Cohen's $d$ , $v$ = Cramér's $v$ , DFA = Discriminant function analysis for variables with small or larger effect sizes, with salient loadings in bold. Effect size (ES): Yellow = small, pink = medium, red = large, -- = not applicable.

People in Class 2, compared to those in Class 1, tended to be younger and perceived themselves as being more affluent (Table 5). There were more Americans in Class 2 (77%) than in Class 1 (49%); that is, more Americans than Canadians tended to be non-adherent to SDIS. People in Class 2 were more likely to believe that the COVID-19 threat was a hoax or exaggerated, even though these individuals were more likely to have reportedly contracted the SARSCOV2 virus. The latter finding is consistent with reports that SARSCOV2 often produces mild illness, particularly among young and healthy individuals (18). People in Class 2 reported greater stressors related to COVID-19 and also reported greater efforts at attempting to cope with those stressors.

## Discussion

Pandemic fatigue is an important problem for managing COVID-19 (1) and likely to be a salient obstacle in mitigating future pandemics. The present study, conducted during the second year of the COVID-19 pandemic, found that past-year decline in adherence to SDIS had a categorical latent structure, consisting of an SDIS adherent group (Class 1: 92% of the sample) and a group reporting a progressive decline in adherence to SDIS (Class 2: 8% of the sample). Class 2 had features indicative of pandemic fatigue; specifically, in addition to reporting a decline in adherence to SDIS, this group had various features consistent with pandemic-related burnout. Compared to Class 1, Class 2 had greater levels of emotional burnout, pessimism, apathy, and cynical or negative beliefs about the COVID-19 pandemic (e.g., believing COVID-19 to be a hoax). The present study confirmed previous findings that pandemic fatigue is associated with the perception that lockdown is unnecessary and ineffective (11), younger age (12-15), greater perceived personal affluence (16), and lower trust in government (16). People in Class 2, compared to Class 1, tended to be more narcissistic, entitled, and gregarious, and were more likely to report having been infected with SARSCOV2, which they regarded as an exaggerated threat. In other words, pandemic fatigue was associated with heightened self-interest to the expense of community needs.

People in Class 2 also reported higher levels of pandemic-related stress, anxiety, and depression, and described making active efforts at coping with SDIS restrictions that they perceived as unnecessary and stressful. People in Class 1 generally reported that they engaged in SDIS for the benefit of themselves and their community, although 35% also feared they would be publicly shamed if they did not comply with SDIS guidelines. The findings suggest that pandemic fatigue affects a substantial minority of people and, importantly, that many SDIS-adherent people also experience emotionally adverse effects (i.e., fear of being shamed) related to SDIS.

Knowledge alone is not enough to overcome pandemic fatigue, as previous surveys have demonstrated that most people are knowledgeable about COVID-19 protective behaviors (1). Consistent with previous surveys (1), we found that the majority of respondents were adherent to SDIS guidelines. During the COVID-19 pandemic, community leaders expressed frustration and dismay at people violating social distancing guidelines, using pejoratives such as “Covidiot” to label these individuals (40). Although public shaming has long played a role in the regulation of societies and can effectively inhibit some forms of socially disruptive behavior (41), public shaming during a pandemic adds a layer of stress on an already distressed public. The burden of accumulated adversity—the piling up of stressors on an individual—is a risk factor for stress-related disorders such as posttraumatic stress disorder (42). Accordingly, shaming people who are already pandemic fatigued—experiencing dysphoria, anxiety, and irritability about COVID-19—is likely to worsen their mental health. Community leaders and others in positions of authority are advised to use caution when considering shaming people who are not complying with SDIS or other pandemic mitigation guidelines.

The present study had various strengths and limitations. In terms of strengths, the sample size was large, robust statistical methods were used, and the assessment period was timely, given that pandemic-related restrictions had been in place for over a year. Regarding limitations, the assessment of SDIS was retrospective, based on self-report, and the generalizability of the results across different demographic and geographic groups remains to be investigated. Retrospective and prospective assessments each have their strengths and limitations, and ideally both would be conducted; but, this was not possible for logistic reasons. Research suggests that behavioral and self-report measures of SDIS produce broadly similar results (6).

Participants were asked to report on their socially undesirable behaviors (i.e., non-adherence to SDIS) and the question arises as to whether the results were affected by a social desirability bias; that is, the tendency to give socially desirable answers to the assessment battery. It might be argued that Class 2 simply represents a group of people who are more willing to admit to socially undesirable attitudes or acts, such as non-adherence to SDIS. This explanation is unlikely for two reasons. First, responding was anonymous. Second, our previous COVID-19 research found that social desirability was unrelated to a range of behavioral, attitudinal, and affective variables (20). Social desirability, as assessed by the Marlowe-Crowne Social Desirability Scale Short Form (43), was uncorrelated (i.e., effect sizes below the threshold for “small”) with scores on various pandemic-related attitudes and behaviors, including disregard for SDIS (*r*=0.03), self-reported violation of pandemic lockdown (i.e., remain-at-home orders) during early 2020 (*r*=0.02), belief that the COVID-19 threat is exaggerated (*r*=-0.01), belief in COVID-19 conspiracy theories (*r*=-0.04), and anti-vaccination attitudes in general (*r*=-0.03) (*N*s ranged from 3,314 to 6,854) (23). Thus, it is unlikely that social desirability affected the results of the present study.

Whether the results of the present study generalize to more protracted, highly restrictive SDIS programs, such as stay-at-home mandates imposed over extended periods of time, remains to be investigated. Under such conditions, the structure of pandemic fatigue, as identified in the present study, may be altered. Non-adherence (as in Class 2) is likely to be found under conditions of more severe lockdowns, unless there are efforts to offset the problem. Pandemic fatigue may also start to appear in people who have been generally adherent (as in Class 1). Future research is also needed to investigate potentially relevant variables that were not examined in the present study. For example, boredom proneness is a trait characterized by the tendency to readily become bored in a wide range of situations (44). This trait was associated with non-adherence to SDIS early in the COVID-19 pandemic and may play a role in pandemic fatigue (45).

Finally, additional research is needed to identify strategies for easing the mental health burden imposed by SDIS. Several studies have found that SDIS harms mental health, with protracted SDIS being correlated with substantial increases in anxiety, depression, substance abuse, and other psychological problems (2). Humans are inherently social creatures, and SDIS involves thwarting this natural urge to socialize. Moreover, research suggests that narcissism (a feature of Class 2 in the present study) is becoming more prevalent in Western societies, likely due to a range of sociocultural factors (46). This raises concerns about the future of pandemic mitigation methods such as SDIS, which require people to work for the collective good rather than focusing on individuals needs or desires.

The WHO described a number of methods intended to reinvigorate people to follow SDIS guidelines (see refs. 1 and 47-49). The efficacy of such methods remains to be established. Encouraging or “nudging” people to follow the guidelines may have greater impact on people who are already amendable to following SDIS guidelines (i.e., Class 1). For people who are narcissistic and distressed, and who see the pandemic restrictions as unnecessary, nudges may be ineffective. Indeed, during the 1918 Spanish flu and during COVID-19, governments responded to non-adherent individuals by becoming increasingly punitive, such as imposing fines or even arresting people who do not comply with SDIS mandates (2, 45). Alternatives to SDIS have been considered, such as the controversial Great Barrington Declaration, which advocates that only the elderly and medically compromised should be subject to stay-at-home orders during COVID-19 (50). This proposal has been widely criticized as discriminatory and likely to result in greater morbidity and mortality than existing SDIS measures (51, 52). During COVID-19, communities experimented with alternatives such as short-term “circuit breaker” lockdowns, in which lockdown and sometimes curfews were imposed for short periods (e.g., two weeks) to attempt to disrupt the spread of infection. The tolerability and efficacy of this and other alternative methods of SDIS that may have less of an impact on mental health remain to be investigated.

## Data Availability

All data produced in the present study are available upon reasonable request to the authors.

## Acknowledgements

The authors thank Michelle M. Paluszek and Caeleigh A. Landry for their assistance in this study.

